# Psychometric analysis of the modified Covid-19 Yorkshire Rehabilitation Scale (C19-YRSm) in a prospective multicentre study

**DOI:** 10.1101/2023.12.22.23300424

**Authors:** Adam B. Smith, Darren C. Greenwood, Mike Horton, Thomas Osborne, Madeline Goodwin, Román Rocha Lawrence, Darren Winch, Paul Williams, Ruairidh Milne, the LOCOMOTION consortium, Manoj Sivan

## Abstract

**Bckground:** Long COVID is a novel multisystem clinical syndrome affecting millions of individuals worldwide. The modified COVID-19 Yorkshire Rehabilitation Scale (C19-YRSm) is a condition-specific patient-reported outcome measure designed for assessment and monitoring of people with Long COVID (LC).

**Objectives:** To evaluate the psychometric properties of the C19-YRSm in a prospective sample of people with Long COVID.

**Methods:** 1314 patients attending UK specialist Long COVID clinics completed C19-YRSm and EQ-5D-5L longitudinally. Scale characteristics were derived for C19-YRSm subscales (Symptom Severity, SS; Functional Disability, FD; and Overall Health, OH) and internal consistency (Cronbach’s alpha). Convergent validity was assessed using the FACIT-Fatigue scale. Known groups validity was assessed for the Other Symptoms (OS) subscale as tertiles, hospitalisation and intensive care admission. Responsiveness and test-retest reliability was evaluated for C19-YRSm subscales and EQ-5D-5L. The minimal important difference (MID) and minimal clinically important difference (MCID) were estimated. Confirmatory factor analysis was applied to determine the instrument’s two-factor structure.

**Results:** C19-YRSm demonstrated good scale characteristic properties. Item-total correlations were between 0.37 to 0.65 (for SS and FD), with good internal reliability (Cronbach’s alphas >0.8). Item correlations between subscales ranged between 0.46 to 0.72. Convergent validity with FACIT was good (−0.46 to −0.62). The three subscales discriminated between different levels of symptom burden (p<0.001), and between patients admitted to hospital and intensive care. There was moderate responsiveness for the three subscales ranging from 0.22 (OH) to 0.50 (SS) and was greater than the EQ-5D-5L. Test-retest reliability was good for both SS 0.86 and FD 0.78. MID was 2 for SS, 2 for FD, and 1 for OH; MCID was 4 for both the SS and FD. The factor analysis supported the two-factor SS and FD structure.

**Conclusions:** The C19-YRSm is a condition-specific, reliable, valid, and responsive patient-reported outcome measure for Long COVID.

**Key messages:** *What is already known on this topic:* Long Covid or Post-COVID-19 syndrome is a multisystem, fluctuating condition. C19-YRSm is literature’s first condition-specific patient reported outcome measure which needed validation in a large population sample.

*What this study adds:* C19-YRSm is a valid, reliable, responsive and easy to administer measure which is able to show clinically meaningful change in the status of the condition in people living with Long Covid.

*How this study might affect research, practice or policy:* C19-YRSm can be used in clinical and research settings to reliably capture the condition trajectory and the effect of interventions and also help inform clinical policy.

## Introduction

Long Covid (LC) or post-acute sequelae of COVID-19 is a fluctuating, multisystem syndrome ^1^ with an estimated prevalence of 1.9 million cases in the UK alone ^2^ and what is estimated to be at least 100 million individuals worldwide ^3^. There have been more than 200 symptoms recorded in LC affecting 10 organ systems. The most commonly reported symptoms include fatigue, cognitive problems, pain, sleep problems, and breathlessness ^4^. These symptoms may persist for extensive periods following the initial COVID-19 infection ^5^. This protracted course of LC leads to a significant negative impact on the individual, in terms of the persistent nature of symptoms and the associated functional disability and adverse health-related quality of life ^2,6^.

The COVID-19 Yorkshire Rehabilitation Scale (C19-YRS) is a condition-specific patient-reported outcome measure designed to capture the symptoms of LC, as well as assess severity and monitor the persistence of symptoms to inform and guide the rehabilitation of affected patients ^7–10^. Since the initial validation of the instrument ^7,11^, it has been widely used in a variety of LC contexts including symptom evaluation in primary care and community settings ^12–14^, determining the need for LC rehabilitation interventions ^8,9,15^, as well as epidemiological assessments of post-COVID symptoms^16,17^.

The original 22-item C19-YRS underwent psychometric evaluation including both classical and modern psychometric evaluation methods resulting in a 17-item modified tool, the C19-YRSm ^18^. Both the original 22-item and the modified version have undergone a limited degree of subsequent validation ^10,19^. The C19-YRS has been shown to have good construct validity but moderate responsiveness ^10^. The C19-YRSm has, by contrast, been demonstrated in a Croatian patient population to have good internal reliability and convergent validity ^19^. Therefore, the aims of this study were to further validate the C19-YRSm with a longitudinal sample of LC patients, as well as to identify minimally (clinically) important differences to inform use in future randomised controlled trials and clinical practice.

## Methods

### Data

The data were collated from the LOng COvid Multidisciplinary consortium Optimising Treatments and servIces acrOss the NHS (LOCOMOTION) study ^20^. This was a prospective mixed-methods study involving 10 LC services across the UK. Ethics approval for the LOCOMOTION study was obtained from the Bradford and Leeds Research Ethics Committee on behalf of Health Research Authority and Health and Care Research Wales (reference: 21/YH/0276) ^20^.

### Participants

Eligibility criteria for the study comprised participants: with a diagnosis of LC who were receiving treatment and management of the condition from one of the 10 participating LC services. Participation in the study required participants to be registered on ELAROS: a digital patient-reported outcome measures platform ^21^. Informed consent and study data were collected on this platform. Participants were requested to complete the following patient-reported outcome measures (see below) every 3 months after registration. The first patient was registered on the 23 November 2021 and the last on 12 November 2023.

### Patient and Public Involvement

Patients have been involved from the outset in the design and implementation of the LOCOMOTION study ^20^ as part of a 9-member Patient Advisory Group (PAG). The PAG has provided the LOCOMOTION study team with first-hand experience of people living with LC. Two co-authors (RM and DW are members of the LOCOMOTION PAG).

## Instruments

### Covid-19 Yorkshire Rehabilitation Scale – Modified (C19-YRSm)

The C19-YRSm is a 17-item instrument ^18^ designed to capture the key symptoms of LC and its impact on activities of daily living and overall health. The items comprise four subscales: Symptom Severity (SS, 10 items), Functional Disability (FD, 5 items), Overall Health (OH, a single item), and Other Symptoms (OS).

The items in the SS subscale comprise the following domains: breathlessness (4 items), cough/throat sensitivity/voice change (2 items), fatigue (one item), smell / taste (2 items), pain / discomfort (five items), cognition (three items), palpitations / dizziness (two items), post-exertional malaise (one item), anxiety / mood (five items), and sleep (one item). FD consists of 5 single items: communication, walking / moving around, personal care, other activities of daily living, and social role.

Responses on the SS and FD subscales are rated on a 0 (no symptom or dysfunction) to 3 (severe life-disturbing symptom or dysfunction) Likert scale. For the SS subscale, the highest value within each of the domains (e.g., breathlessness, pain / discomfort) are added to determine the score for that subscale. Higher scores on both these subscales indicate worse symptomatology and poorer functioning. Responses on the OH subscale are scored on a 0-10 Likert scale (0 being “worst health” and 10 being “best health”) with higher scores indicating better health. Other Symptoms (OS) over the last 7 days are also captured from a list of 25 additional symptoms ^18^.

### Functional Assessment of Chronic Illness Therapy – Fatigue Scale (FACIT-Fatigue)

The FACIT-Fatigue scale is a 13-item instrument developed to evaluate fatigue and its impact on health-related quality of life and daily activities ^22^. Responses are scored on a 5-item Likert scale (from 0 to 4) with a maximum total score of 52 (range 0 to 52). Higher scores indicate better health-related quality of life. Items in the FACIT-Fatigue cover tiredness, fatigue, listlessness, lack of energy, and the impact on daily and social activities. Although originally developed for use with cancer patients, the FACIT-Fatigue scale has been used to evaluate post-COVID fatigue ^23^.

### The EuroQol 5D-5L (EQ-5D-5L)

The EuroQol EQ-5D-5L is a preference-based instrument with five domains: Mobility, Usual Activities, Selfcare, Pain / Discomfort, and Anxiety / Depression ^24^. It has five response categories ranging from 1 (no problems) to 5 (severe problems). Responses to each domain are collated into a profile score which is converted into a health utility or index score using a country-specific algorithm (tariff or value set). Utilities reflect societal preferences for health states and are measured on a metric from 0 (dead) to 1 (perfect health). Utility values less than 0, indicating states worse than dead, are also captured. The EQ-5D-5L scores were mapped onto the EQ-5D-3L using the crosswalk (CW) algorithm to derive UK utility values ^25^. The EQ-5D also comprises a visual analogue scale (VAS) measuring self-reported current health on a scale from 0 (“worst health”) to 100 (“best health”).

## Statistical analysis

All analyses were performed using R Studio (R version 4.1.1). Descriptive summary statistics (mean, standard deviation, count and percentage) were generated for the following patient demographic and clinical data: age, sex, ethnicity, smoking status, hospital admission, intensive care (ICU) admission. The scale characteristics for the three C19-YRSm subscales (SS, FD, and OH) were derived including: mean (standard deviation, SD), median (inter-quartile range), score range, and skewness (evaluated −0.5 to +0.5). Item characteristics, such as mean item score (SD), missing values, floor and ceiling effects, and item-total correlations were estimated for the SS and FD domains.

The internal reliability of the C19-YRSm SS and FD domains was evaluated using Cronbach’s alpha. A Cronbach’s alpha >0.7 was considered to be an indicator of adequate internal consistency and >0.8 was considered to be an indicator of good internal consistency.

Convergent validity – the degree to which items or domains on different instruments measure the same constructs – was assessed for the C19-YRSm subscales, SS, FD, and OH using the FACIT-Fatigue. As both SS and FD are negatively scored (a higher score indicates worse symptoms or functioning), negative associations were anticipated between these and the FACIT-Fatigue score. Conversely, positive associations were hypothesised between OH and FACIT-Fatigue. Associations were evaluated using Pearson’s product moment.

Known-groups validity was assessed for the C19-YRSm domains using the OS subscale split into tertiles: low number of symptoms (0-3), medium number of symptoms (4-7), and high number of symptoms (7+), as well as hospitalisation and admission to ICU (yes/no). Kruskall-Wallis and Mann-Whitney-Wilcoxon rank sum tests were used to evaluate differences in scores across these pre-defined groups.

Responsiveness of the three C19-YRSm subscales (SS, FD, and OH) were evaluated using a subset of patients who had completed the instrument at two timepoints, namely the first assessment and at follow-up 30 days later (± 10 days). The responsiveness of the EQ-5D-5L and EQ-5D VAS was also evaluated as a comparator for those patients who had completed both the C19-YRSm and EQ-5D-5L on the same day (at first assessment and 30 days (± 10 days)).

Mean change from the first assessment was derived for these domains and an effect size was calculated (standardised mean response, SMR) by dividing this by the standard deviation of the mean change scores. Intra-class correlations and test-retest reliability were also derived to evaluate stability in the instrument subscales over time. Test-retest reliability was evaluated against OH: the reliability coefficient was derived for patients with no change score on the OH between first assessment and day 30 (± 10 days).

A half standard deviation of the first assessment domain scores was applied as a putative minimally important difference (MID) [26]. In addition to this, the standard error of measurement (SEM) and reliable change index (RIC) were calculated as follows for the SS and FD domains as indicators of minimal clinically important differences (MCIDs):

SEM = 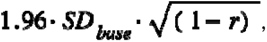, and r is the test-retest reliability coefficient.

RIC = 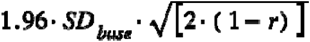, where SD_base_ is the standard deviation at first assessment.

The putative factor structure – a two-dimensional structure encompassing the SS and FD domains – was explored utilising a confirmatory factor analysis (CFA). A number of indices were employed to evaluate the goodness-of-fit of the model: root mean square error of approximation (RMSEA) ^27^, comparative fit index (CFI) ^28^, the Tucker-Lewis index (TLI) ^29,30^, and the standardised root mean squared residual (SRMR) ^31^. Various thresholds have been proposed to evaluate model fit. In this study, RMSEA <0.08 ^32^ was considered to be a reasonable fit; TLI and CFI >0.90 as acceptable fit ^29^, and SRMR <0.08 as acceptable fit ^31^. As no single index provides sufficient evidence alone of model fit, four indices were evaluated in aggregate. The *lavaan* package in R was used for the CFA.

## Results

### Demographics

A total of 1314 patients (Table 1) had completed the C19-YRSm on at least one occasion; 263 patients (20%) had completed the instrument at first assessment and at 30 days (± 10 days), and 193 patients had completed the FACIT-Fatigue instrument at least once (15%). The C19-YRSm and EQ-5D-5L had been completed on the same day at both timepoints (first assessment and day 30 (± 10 days)) by 98 patients. The majority (total sample) were Caucasian (76%) females (67%) with an average age of 48 years (SD: 13 years); 10% had been admitted to hospital as a result of COVID-19, and just over 2% had been admitted to ICU.

**Table 1.** Basic sociodemographic details.

| Characteristic | N = 1,314<sup>I</sup> |
| --- | --- |
| **Sex** |  |
| Female | 882 (67%) |
| Male | 432 (33%) |
| **Age (years)** | 47.9 (13.0) |
| **Ethnicity** |  |
| Asian (includes any Asian background, for example, Bangladeshi, Chinese, Indian, Pakistani) | 84 (6.4%) |
| Black, African, black British or Caribbean (includes any black background) | 29 (2.2%) |
| Mixed or multiple ethnic groups (includes any mixed background) | 20 (1.9%) |
| White (includes any white background) | 1003 (76%) |
| Another ethnic group (includes any other ethnic group, for example, Arab) | 18 (1.4%) |
| Not recorded | 152 (12%) |
| **Smoking status** |  |
| Current occasional smoker | 21 (1.6%) |
| Current regular smoker | 11 (0.8%) |
| Ex-smoker | 105 (8%) |
| Never smoked | 271 (21%) |
| Not recorded | 906 (69%) |
| **Hospital admission** |  |
| No | 1179 (90%) |
| Yes | 134 (10%) |
| **ICU admission** |  |
| No | 1283 (98%) |
| Yes | 31 (2.0%) |

<sup>I</sup> N (%)

The mean subscale scores are shown in Table 2. Both means for the SS (18.4, SD: 5.62) and FD (7.1, SD: 3.78) subscales suggested a moderate-to-high level of symptom burden and functional disability. Similarly, OH indicated that patients were at best in moderate health. These three subscales showed little skewness reflecting symmetrical score distributions. The mean number of OS was 5 with positive skew (fewer patients with large numbers of other symptoms).

**Table 2.** C19-YRSm domain characteristics.

| **Subscale (scale range)** | **Valid scores (N)** | **Mean (SD)** | **Median (IQR)** | **Score range** | **Skewness** |
| --- | --- | --- | --- | --- | --- |
| **Symptom Severity (0-30)** | 1314 | 18.4 (5.62) | 18 (14.5 - 22) | 0 - 30 | -0.17 |
| **Functional Disability (0-15)** | 1314 | 7.1 (3.80) | 7.0 (4.0 - 10.0) | 0 - 15 | 0.13 |
| **Overall Health (0-10)** | 1314 | 4.5 (1.90) | 5.0 (3.0 - 6.0) | 0 - 10 | 0.13 |
| **Other Symptoms (0-25)** | 1314 | 5.6 (4.3) | 5.0 (2.0 – 8.0) | 0 – 23 | 0.78 |

\*N, number; SD, standard deviation; IQR, interquartile range

The item means (Table 3) for the SS and FD subscales ranged approximately between 1 and 2 indicating that patients were on average experiencing at least mild (to moderate) symptom burden and functional disability, although this varied across the items as reflected in the results of the floor and ceiling effects. Missing data was negligible (<3%).

**Table 3.**
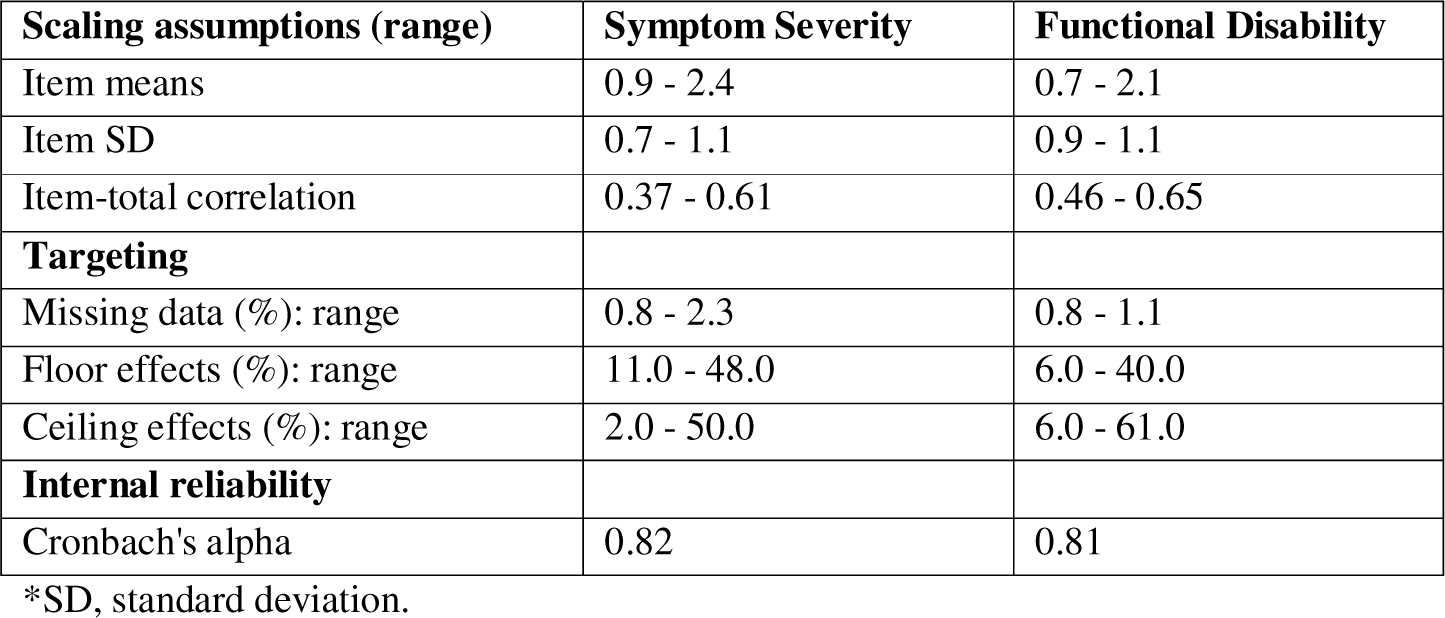
Scaling assumptions, targeting, and internal reliability.

| **Scaling assumptions (range)** | **Symptom Severity** | **Functional Disability** |
| --- | --- | --- |
| Item means | 0.9 - 2.4 | 0.7 - 2.1 |
| Item SD | 0.7 - 1.1 | 0.9 - 1.1 |
| Item-total correlation | 0.37 - 0.61 | 0.46 - 0.65 |
| **Targeting** |  |  |
| Missing data (%): range | 0.8 - 2.3 | 0.8 - 1.1 |
| Floor effects (%): range | 11.0 - 48.0 | 6.0 - 40.0 |
| Ceiling effects (%): range | 2.0 - 50.0 | 6.0 - 61.0 |
| **Internal reliability** |  |  |
| Cronbach's alpha | 0.82 | 0.81 |

\*SD, standard deviation.

### Convergent validity

Table 4 shows the correlation matrix between the domains (see also Supplementary Figure 1). There was a strong positive association between SS and FD. A moderate positive association was determined between OS and SS, and OS and FD. OH was negatively associated with the SS, FD, and OS.

**Table 4.** Correlation matrix C19-YRSm domains.

| **Domains** | **Symptom Severity** | **Functional Disability** | **Overall Health** | **Other Symptoms** |
| --- | --- | --- | --- | --- |
| **Symptom Severity** | 1.00 | 0.73 | -0.47 | 0.63 |
| **Functional Disability** | 0.73 | 1.00 | -0.52 | 0.55 |
| **Overall Health** | -0.47 | -0.52 | 1.00 | -0.34 |
| **Other Symptoms** | 0.63 | 0.55 | -0.34 | 1.00 |

\*N=1314; p<0.001

There was a strong negative association between the total FACIT score and the C19-YRSm Fatigue item (r = −0.58, p<0.001, 95%CI: −0.67 to −0.48), and similarly for SS (r = −0.61, p<0.001, 95%CI: - 0.69 to −0.51), FD (r = −0.64, p<0.001, 95%CI: −0.72 to −0.55), and OS (r = −0.46, p<0.001, 95%CI: - 0.56 to −0.34). The FACIT-Fatigue total was positively associated with OH (r = 0.47, p<0.001, 95%CI: 0.36 to 0.58).

### Known-groups validity

There was a linear increase in SS score as symptom burden (tertiles of the OS domain) increased in severity from low to high (Table 5a). A similar pattern was observed for FD, whereas OH showed a decrease as symptom burden increased. All these results were statistically significant (p<0.001).

**Table 5.** Responsiveness C19-YRSm domains.

| **Subscale (scale range) /**<br>**Change from 1st assessment** | **Mean (SD)** | **Median** | **Min, Max** | **ICC** | **TRR** | **ES** | **MID** | **MCID**<br>**(SEM)** | **MCID**<br>**(RIC)** |
| --- | --- | --- | --- | --- | --- | --- | --- | --- | --- |
| **Symptom Severity (0-30)** | 2.0 (4.0) | 2 (0 to 3) | (-19.0, 23.0) | 0.68 | 0.86 | 0.50 | 2 | 4 | 6 |
| **Functional Disability (0-15)** | 0.8 (2.4) | 1 (-1 to 2) | (-5.0, 10.0) | 0.76 | 0.78 | 0.33 | 2 | 4 | 5 |
| **Overall Health (0-10)** | 0.3 (1.67) | 0.4 (-1 to 1) | (-7.0, 7.0) | 0.58 | - | 0.22 | 1 | - | - |

\*SD, standard deviation; min, minimum; max, maximum; ICC, intraclass correlation; TRR, test-retest reliability; ES, effect size; MID, minimally important difference; MCID, minimal clinically important difference; SEM, standard error of measurement; RIC, reliable change index

Patients who had been hospitalised for COVID-19 showed higher SS and FD scores (Supplementary Tables 1a and 1b) compared to those who had not been hospitalised (p=0.04 and p=0.008, respectively). No differences between these groups were observed for OH. Statistically significant differences for both SS and FD (but not OH) were also observed between those who had and had not been admitted to ICU (Supplementary Tables 1c).

### Responsiveness

The mean change over 30 days (± 10 days) was 1.9 (SD: 4.38) for the SS domain. Smaller changes were observed in both the FD and OH domains, 0.7 (SD: 2.53) and 0.3 (SD: 1.67) (Table 5).

The intraclass correlation coefficients for the 3 domains ranged from 0.58 (OH) to 0.76 (FD) (Table 5) suggesting moderate-to-strong content structure over time. A total of 70 patients had stable (unchanged) OH scores over the 30-day evaluation period (± 10 days). The test-retest reliability coefficient for the SS domain was 0.86 and 0.78 for the FD domain, indicating good reliability.

All 3 subscales demonstrated a degree of responsive to change (effect sizes range: 0.22 to 0.50) (Table 5). The responsiveness of the EQ-5D-5L Index was by comparison 0.14 and 0.18 for the VAS.

The 0.5 SD was applied as a metric for the MID. This resulted in the following MIDs: SS = 2; FD = 2; and OH = 1. From Table 6, it may be seen, for instance, that a MID was recorded for SS over the 30- day period following first assessment, but not for either FD or OH. The MCID estimate (based on the SEM) was 4 for both the SS and FD (Table 5).

### Factor Structure

The results of the confirmatory factor analysis showed a RMSEA of 0.10 (90% confidence intervals, CI: 0.096, 0.107) (Figure 1). The SRMR was 0.066, CFI 0.83, and Tucker-Lewis Index 0.8. Taking all 4 indices together, these indicated reasonable model fit for the two-factor model. These factors were consistent with the interpretation of one factor measuring SS and the other measuring FD.

**Figure 1.**
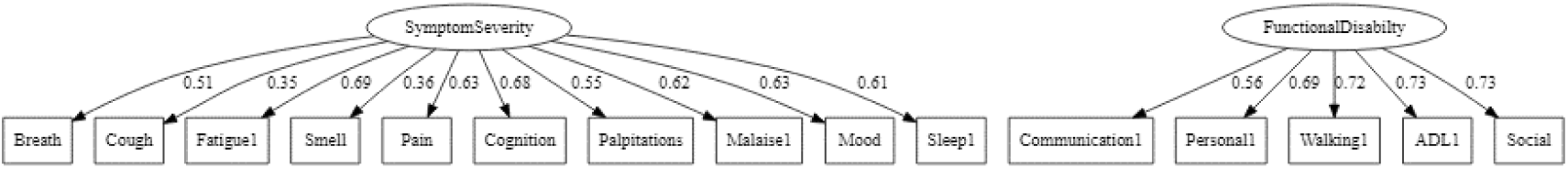
Factor structure of the C19-YRSm Symptom Severity and Functional Disability Domains.

## Discussion

The aim of this study was to undertake a further psychometric validation of the C19-YRSm. The results demonstrated good item and scale characteristics. There was good convergent validity with the FACIT-Fatigue scale. Furthermore, the three subscales (SS, FD, and OH) discriminated well between levels of symptom severity, and between patients who had been hospitalised and admitted to ICU. There was also good internal reliability, test-retest reliability, and stability of the subscale scores over time. Furthermore, the convergent correlations were as hypothesised.

The results of the responsiveness analysis showed that the instrument was able to detect changes as patients’ symptoms fluctuated and was more sensitive to change than the generic health-related quality of life measure, the EQ-5D-5L (both Index and VAS). This is a potentially important finding for future randomised controlled trials in LC. Although the effect sizes were modest, these must be evaluated in the context of a fluctuating condition ^4, 33–34^, and it may therefore be that potentially larger effect sizes were being masked by frequent changes in symptoms. The results also suggest some initial metrics for the MID for SS (2), FD (2), and OH (1), as well as the MCID (4 SS and FD). Although these were based on distribution methods, and therefore remain to be confirmed using anchor-based approaches such as patient and clinician global impression of change, these provide useful initial metrics for interpreting meaningful changes in the C19-YRSm scores to aid both clinical interpretation as well as inform sample size considerations for prospective randomised controlled trials.

There was also some support for a 2-factor structure, although the statistics in isolation did not meet the pre-defined thresholds. Although there are no definitive guidelines on what constitutes ideal fit, it is possible that model fit could be improved to a degree that may consequently also positively impact on responsiveness. Previous research has similarly determined moderate responsiveness at the item-level ^10^. Further research involving modern psychometric analysis such as Rasch or Item-response theory could explore this issue further and potentially identify individual items that may be removed and / or recalibrated to improve instrument responsiveness.

The results of this study are in line with a previous psychometric validation study of the C19-YRSm, which found both good internal reliability and convergent validity of the instrument ^19^, providing further evidence for the psychometric properties of the C19-YRSm with meaningful factors or domains, such as SS, FD, and OH. The latter is further bolstered by the large sample size in this study and builds on the earlier development of the instrument ^18^, supporting its use as one of the few LC-specific patient-reported outcome measures. In addition, it is shorter than other condition-specific instruments such as the Symptom Burden Questionnaire for Long COVID with 131 items ^35^, thereby minimising patient burden – a particularly important factor in people living with LC, who may present with fatigue and cognitive dysfunction. The instrument’s brevity and design lends its use for self-completion by patients, enabling the fluctuating nature of the condition to be monitored on a frequent basis for patients’ own awareness, for instance, in determining symptom triggers, as well as by clinicians to evaluate patients’ condition over time between clinics appointments.

Given the prevalence of LC, its associated persistence of debilitating symptoms, and the impact of the condition on patients’ health-related quality of life, valid and reliable condition-specific patient-reported instruments such as the C19-YRSm are of critical importance in the assessment of LC symptoms, as well as in helping to facilitate appropriate management and rehabilitation of patients suffering with the condition. The evidence presented alongside other studies ^19^ suggest that C19-YRSm is a condition-specific, reliable, valid, and responsive patient-reported outcome measure for Long COVID.

## Data Availability

All data produced in the present work are contained in the manuscript

---

**The C19-YRSm is available at:** https://c19-yrs.com/the-c19-yrs/

**Supplementary Table 1a.** C19-YRSm domain mean domain scores by symptom burden (“Other Symptoms”)

| Domain | Low, N = 507<sup>1</sup> | Medium, N = 441<sup>1</sup> | High, N = 396<sup>1</sup> | p-value<sup>2</sup> |
| --- | --- | --- | --- | --- |
| **Symptom Severity (0-30)** | 14.6 (4.8) | 18.8 (4.5) | 22.8 (4.2) | <0.001 |
| **Functional Disability (0-15)** | 5.1 (3.1) | 7.0 (3.3) | 9.9 (3.3) | <0.001 |
| **Overall Health (0-10)** | 5.1 (1.8) | 4.4 (1.7) | 3.8 (1.8) | <0.001 |

<sup>1</sup>Mean (Standard Deviation); <sup>2</sup>Kruskal-Wallis rank sum test; Low, Medium, High based on tertiles of the “Other Symptom” domain

**Supplementary Table 1b.** C19-YRSm domain mean domain scores by hospital admission.

| Hospital Admission | No, N = 1179<sup>1</sup> | Yes, N = 134<sup>1</sup> | p-value<sup>2</sup> |
| --- | --- | --- | --- |
| **Symptom Severity (0-30)** | 18.3 (5.6) | 19.3 (6.0) | 0.07 |
| **Functional Disability (0-15)** | 7.0 (3.7) | 8.0 (4.2) | 0.03 |
| **Overall Health (0-10)** | 5.0 (1.9) | 4.5 (1.9) | 0.8 |

<sup>1</sup>Mean (Standard Deviation); <sup>2</sup>Mann-Whitney-Wilcoxon rank sum test; Missing values, N=8

**Supplementary Table 1c.** C19-YRSm domain mean domain scores by ICU admission.

| Characteristic | No, N = 1,283<sup>1</sup> | Yes, N = 31<sup>1</sup> | p-value<sup>2</sup> |
| --- | --- | --- | --- |
| **Symptom Severity (0-30)** | 18.3 (5.6) | 20.2 (6.0) | 0.07 |
| **Functional Disability (0-15)** | 7.1 (3.8) | 8.8 (4.5) | 0.03 |
| **Overall Health (0-10)** | 4.5 (1.9) | 4.3 (2.2) | 0.9 |

<sup>1</sup>Mean (Standard Deviation); <sup>2</sup>Mann-Whitney-Wilcoxon rank sum test; Missing values, N=8

**Supplementary Figure 1.**
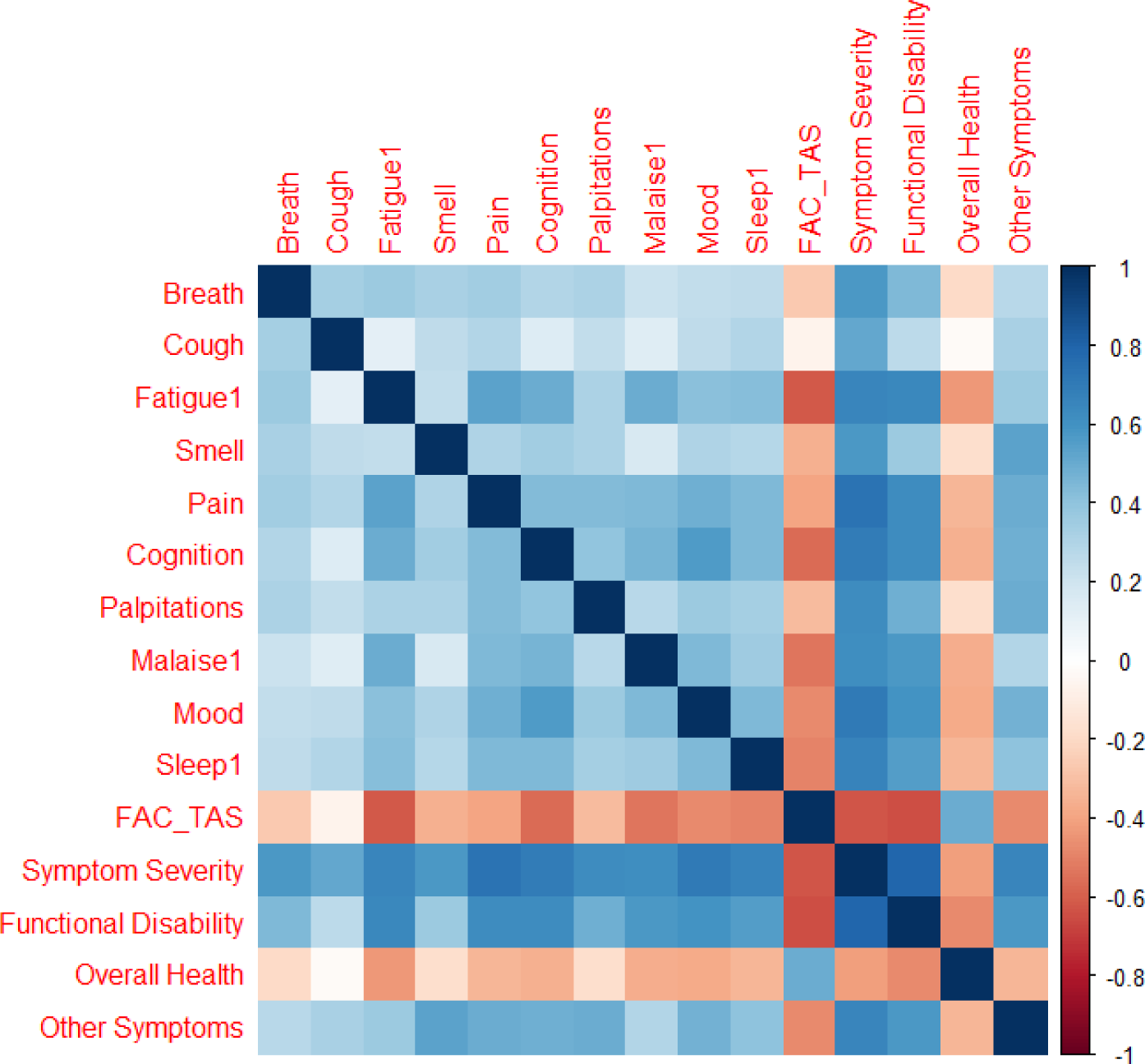
Correlation matrix C19-YRSm and the FACIT total.

